# Complex genetic interactions affect susceptibility to Alzheimer’s disease risk in the *BIN1* and *MS4A6A* loci

**DOI:** 10.1101/2024.10.24.24316008

**Authors:** Alireza Nazarian, Marissa Morado, Alexander M. Kulminski

## Abstract

Genetics is the second strongest risk factor for Alzheimer’s disease (AD) after age. More than 70 loci have been implicated in AD susceptibility so far, and the genetic architecture of AD entails both additive and nonadditive contributions from these loci.

To better understand nonadditive impact of single-nucleotide polymorphisms (SNPs) on AD risk, we examined individual, joint, and interacting (SNPxSNP) effects of 139 and 66 SNPs mapped to the *BIN1* and *MS4A6A* AD-associated loci, respectively. The analyses were conducted by fitting three respective dominant allelic-effect models using data from four independent studies. Joint effects were analyzed by considering pairwise combinations of genotypes of the selected SNPs, i.e., compound genotypes (CompG).

The individual SNP analyses showed associations of 18 *BIN1* SNPs and 4 *MS4A6A* SNPs with AD. We identified 589 *BIN1* and 217 *MS4A6A* SNP pairs associated with AD in the CompG analysis, although their individual SNPs were not linked to AD independently. Notably, 34 *BIN1* and 10 *MS4A6A* SNP pairs exhibited both significant SNPxSNP interaction effects and significant CompG effects. The vast majority of nonadditive effects were captured through the CompG analysis.

These results expand the current understanding of the contributions of the *BIN1* and *MS4A6A* loci to AD susceptibility. The identified nonadditive effects suggest a significant genetic modulation mechanism underlying the genetic heterogeneity of AD in these loci. Our findings highlight the importance of considering nonadditive genetic impact on AD risk beyond the traditional SNPxSNP approximation, as they may uncover critical mechanisms not apparent when examining SNPs individually.

## 1. Introduction

Alzheimer’s disease (AD) is the most common cause of dementia [1], which in most cases has a multifactorial nature [2]. The complex genetic architecture of AD entails both additive and epistatic contributions from multiple loci [3–6]. The *APOE* (Apolipoprotein E) locus on chromosome 19q13.3 harbors the main AD-associated variants (e.g., ε2 and ε4 alleles) [1,7,8]. In addition, there are several genes on the other chromosomal regions whose AD associations have been replicated in multiple independent studies [9,10], including *BIN1* (Bridging Integrator 1) [11–16] and *MS4A6A* (Membrane-Spanning 4-domain subfamily A 6A) [12,17–21], among others.

The *BIN1* gene on chromosome 2q14.3 [22] is deemed as the second most important contributing locus to the genetic architecture of AD after *APOE* [16]. It encodes multiple transcript isoforms, which play roles in processes such as synaptic function, homeostasis, and immune responses [16,22]. Several single-nucleotide polymorphisms (SNPs) in the *BIN1* locus have been associated with AD by genome-wide association studies (GWAS) [10], which are mostly noncoding variants [23], such as rs6733839 [15], rs4663105 [14], rs7561528 [13], and rs744373 [12] [with P=6.00E-118, 4.00E-58, 2.00E-15, and 3.00E-14, respectively]. *BIN1* is among the highly expressed genes in the brain microglia and may contribute to the pathogenesis of AD by regulating microglial inflammatory responses [16]. The overexpression of neuronal isoform of *BIN1* has been linked to endosomal vesicles enlargement [24] and tau-dependent neurodegeneration and neuronal hyperexcitability [16,23,25].

The *MS4A6A* gene is a member of the *MS4A* genes cluster family on chromosome 11q12.2 [22]. The *MS4A* genes encode transmembrane proteins, which may act as G-protein-coupled receptors and be involved in cell signaling, endocytosis, and immune responses, etc. [18,19,22,26]. Some of the *MSA4* genes, such as *MS4A6A* and *MS4A4E* have been implicated in AD [9,10,12,17–19]. For instance, rs367670643 [27], rs11605427 [20], rs2081545 [21], rs610932 [12], and rs7935829 [21] [with P=2.00E-21, 8.00E-17, 1.55E-15, 2.00E-14, and 8.00E-13, respectively] are among the top AD-associated *MS4A6A* variants identified by previous GWAS [10]. The *MS4A6A* and *MS4A4E* genes were found to be highly expressed in immune cells like brain microglia [18,19]. In addition, *PU*.*1* (*SPI1*), which is an AD-associated transcription factor and is highly expressed in immune cells was reported to regulate the expressions of some *MS4A* genes like *MS4A6A* [18]. It has been reported that several AD-associated SNPs within the *MS4A* 11q12.2 locus could modify the concentration of soluble TREM2 (sTREM2) in cerebrospinal fluid (CSF) [28]. The sTREM2 is a marker of microglial activation [19] and its higher CSF level has been associated with delayed age at onset of AD and increased *MS4A6A* expression in brain [19]. The overexpression of *MS4A6A* in the brain has been associated with higher Braak scores [29,30] in AD-affected subjects [18].

The study of complex nonadditive genetic effects (e.g., interactions, combined genotypes, haplotypes) on AD risk has been of great interest in both *APOE* locus [5,6,31–39] and non-*APOE* loci [4,10,40–44]. For instance, we have recently identified genome-wide associations of the *APOE* ε2 and ε4 alleles (encoded by rs7412 and rs429358, respectively) with AD-specific effects (i.e., effect sizes were significantly different between AD-affected and AD-unaffected groups). Such associations could modulate the effects of these alleles on the AD risk [5]. The ε2- and ε4-encoding SNPs were also in AD-specific linkage disequilibrium (LD) with multiple inter- and intra-chromosomal SNPs [39]. As another example, we have identified multiple SNP pairs in the *CLU* (Clusterin) and *ABCA7* (Adenosine triphosphate Binding Cassette subfamily A member 7) loci, which were jointly (i.e., in the form of combinations of genotypes, called compound genotypes) associated with AD risk, while none of their comprising SNPs could modify AD risk individually [43].

Following our previous analyses on the complex AD associations in the *APOE, CLU*, and *ABCA7* loci, we investigated whether such associations may exist for the SNPs mapped to the *BIN1* and *MS4A6A* loci. Thus, 139 and 66 SNPs in these loci were selected, respectively, and their individual and pairwise associations with AD were examined by fitting single SNP, compound genotype, and conventional SNPxSNP interaction models using data from four independent studies. These analyses provided novel insights into complex genetic architecture of AD at these loci. In particular, we found that 34 *BIN1* and 10 *MS4A6A* SNP pairs were associated with AD in both compound genotype and interaction analyses. The SNPs comprising 26 and 9 of these pairs, respectively, were not associated with AD individually

## 2. Methods

### 2.1 Study Participants

Genotypic and phenotypic data from individuals of European ancestry was obtained from three cohorts from the National Institute on Aging (NIA) Alzheimer’s Disease Centers (ADC1-ADC3), which are a part of the Alzheimer’s Disease Genetics Consortium (ADGC) [17], Alzheimer’s Disease Sequencing Project (ADSP-WGS) [45,46], NIA’s Late-Onset Alzheimer’s Disease Family-Based Study (LOAD-FBS) [47,48], and the United Kingdom Biobank (UKB) datasets [49]. Any overlapping subjects between LOAD-FBS and ADSP-WGS were excluded. Furthermore, AD-unaffected individuals of 65 years of age and younger were excluded from the UKB dataset. AD status in the non-UKB datasets was reported by the study researchers according to the National Institute of Neurological and Communicative Disorders and Stroke and the AD and Related Disorders Association (NINCDS-ADRDA) guidelines [48,50,51]. The ICD-10 codes (International Classification of Disease Codes, 10th revision) were used to determine the case-control status of subjects in UKB. A detailed breakdown of study participants’ information by dataset can be seen in Table S1.

### 2.2. Genotype Data and Quality Control

To ensure rigorous quality control, all *BIN1* and *MS4A6A* SNPs in this study met the following criteria. SNPs were selected 500 kb up- and downstream of the *BIN1* locus (i.e., within 500kb of the start and end position of the gene). Accounting for the overlapping nature of *MS4A6A* and *MS4A4E* genes up-/downstream regions (they are located within ∼16 kb of one another), SNPs were selected 500 kb downstream from *MS4A6A* and 500 kb upstream from *MS4A4E*. Furthermore, both *BIN1* and *MS4A6A* SNPs had an imputation quality cutoff of r^2^=0.9 and pairwise LD pruning cutoff of r^2^=0.7. In addition, SNPs with a Hardy-Weinberg equilibrium (HWE) P-value below 1E-6 were excluded from both genes to avoid genotyping errors, and all chosen SNPs had minor allele frequency (MAF) of 5% or larger. SNPs with a missing genotyping rate of 5% or larger were also excluded from this analysis. Ultimately, 139 *BIN1* SNPs and 66 *MS4A6A* SNPs met the above criteria (Table S2). Quality control was performed using *PLINK* (v2.0) (www.cog-genomics.org/plink/2.0/) [52].

### 2.3 Association Analysis of AD Risk

Three separate analyses were performed to expound the complex genetic associations between AD and SNPs in each gene. These included a single SNP analysis, a compound genotype (CompG) analysis for each SNP pair, and a conventional SNPxSNP interaction analysis for each SNP pair. For these analyses, genotypic SNP information utilized binary dominant allelic effect (i.e., 0=major allele homozygote, 1=heterozygote or minor allele homozygote) in lieu of the additive allelic counts to avoid issues with small allelic count sample sizes, particularly minor allele homozygotes. All models were adjusted for age, sex, as well as for the *APOE* ε2 and ε4 variants (SNPs rs7412 and rs429358, respectively) as fixed-effects covariates. The LOAD-FBS dataset was additionally adjusted for a random effect covariate, family-ID, to account for familial clustering. Necessary packages for the association analyses included *stats* (v4.3.2) [53] and *lme4* (v1.1.35.1) [54] packages in R (v4.3.2) [53]. Models with the LOAD-FBS dataset utilized the *glmer* function [54] as it had a random-effect model component that adjusted for familial correlation, while the other three datasets (ADSP-WGS, ADGC, UKB) utilized the *glm* function [53] for their analyses.

In total, 9591 models for the *BIN1* (pairwise combinations of 139 SNPs) and 2145 models for the *MS4A6A* (pairwise combinations of 66 SNPs) were fitted for each respective dataset under the conventional SNPxSNP interaction and CompG analyses. Table 1 defines the four CompG categories constructed for the analysis.

**Table 1.** Compound genotype defined for each SNP pair in our analyses.

| Minor Allelic Count |  | Compound Genotype |
| --- | --- | --- |
| SNP1 | SNP2 |  |
| 0 | 0 | MM |
| 1 or 2 | 0 | mM |
| 0 | 1 or 2 | Mm |
| 1 or 2 | 1 or 2 | mm |

Notations: SNP = single-nucleotide polymorphism.

Here, the reference factor category in our models is represented by the compound genotype MM, the case in which both SNP1 and SNP2 are comprised of major allele homozygotes. In addition, pairwise differences between compound genotype categories (i.e., Mm, mM, and mm) were tested using the chi-square test with one degree of freedom, defined as below [55]:

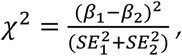

where *β*_1,2_ and SE_1,2_ denote the effect sizes and standard errors of the associations for the two compared compound genotype categories.

The third analysis was a single SNP analysis conducted for each of the 139 SNPs in *BIN1* and 66 SNPs in *MS4A6A* to evaluate their individual association with AD. Here, we also utilized the chi-square test to compare the difference between effect sizes of the individual SNP analysis and the effects of three CompG categories.

### 2.4 Meta-Analysis of AD Risk

Following individual SNP, CompG, and interaction analyses in each dataset under consideration (i.e., ADGC, ADSP-WGS, LOAD-FBS, and UKB), a fixed-effects inverse variance meta-analysis was performed on each model through the *metafor* (v4.4.0) R package [53,56]. To account for the multiple tests, P-values were adjusted through the Sequential Goodness-of-Fit (SGoF) multiple test procedure using the *sgof* (v2.3.4) R package [53,57]. Significant effects and chi-squared differences were identified at the SGoF-adjusted P-values less than 0.05.

## 3. Results

Summary results regarding AD-associated *BIN1* and *MS4A6A* SNPs in the single SNP meta-analyses are provided in Table 2. Also, Tables 3 and 4 contain summary results regarding the AD-associated *BIN1* and *MS4A6A* SNP pairs, respectively, which had both significant CompG effects and significant SNPxSNP interaction terms in the traditional interaction analyses. Detailed results regarding significant findings from CompG and interaction meta-analyses of the *BIN1* SNPs and information regarding their pairwise LD are provided in Tables S3-S6. Detailed results from CompG and interaction meta-analyses and LD analyses of the *MS4A6A* SNPs are provided in Tables S7-S10.

**Table 2.**
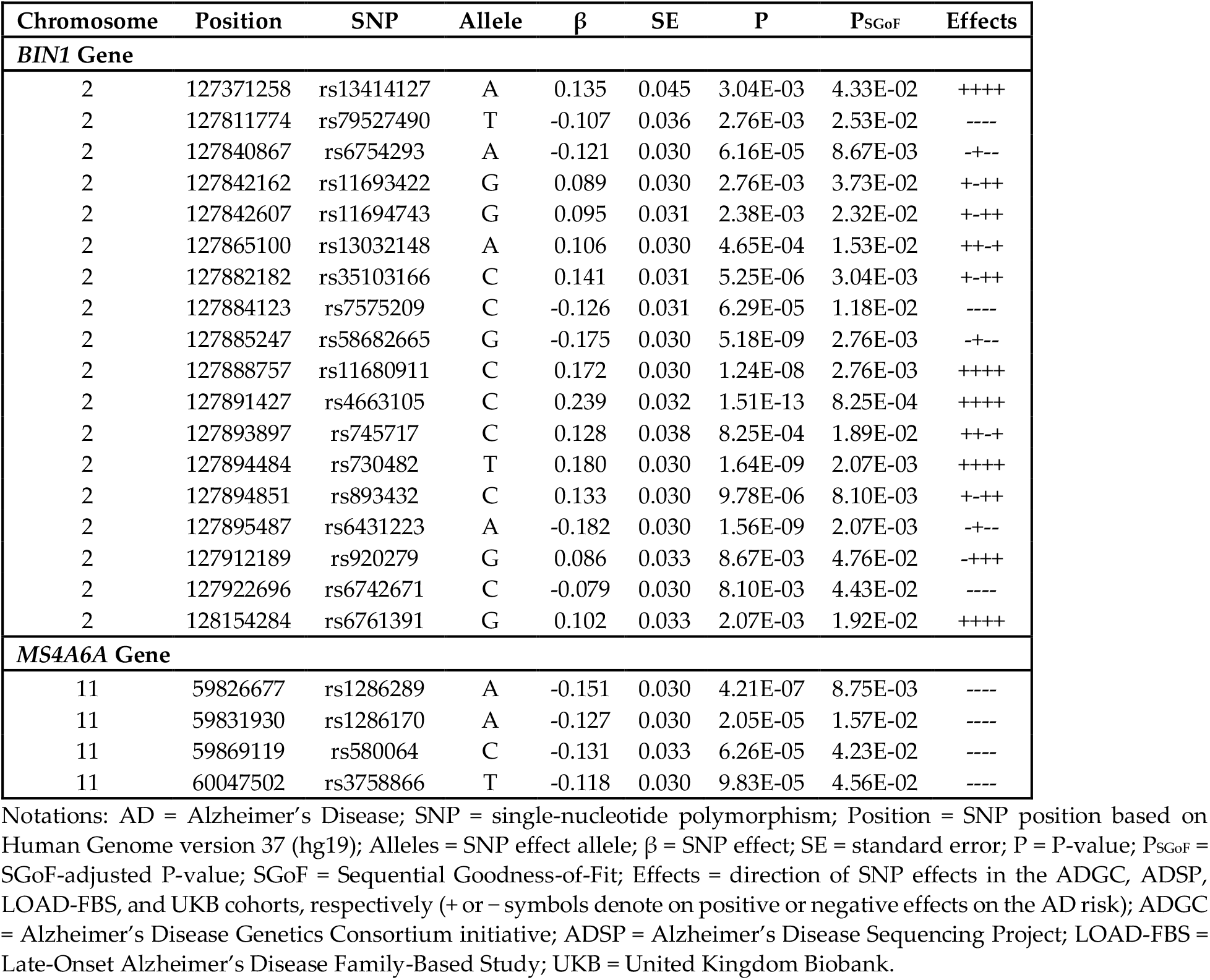
Significant findings from single SNP meta-analyses.

**Table 3.** Thirty-four AD-associated SNP pairs mapped to the *BIN1* gene locus with significant CompG effects and SNP-by-SNP interactions.

| SNP Pair |  | Compound Genotype Model |  |  |  |  |  | Interaction Model |  |  |  |  |  |
| --- | --- | --- | --- | --- | --- | --- | --- | --- | --- | --- | --- | --- | --- |
| SNP1 | SNP2 | Mm |  | mM |  | mm |  | SNP1 |  | SNP2 |  | Int |  |
|  |  | β | P<sub>SGoF</sub> | β | P<sub>SGoF</sub> | β | P<sub>SGoF</sub> | β | P<sub>SGoF</sub> | β | P<sub>SGoF</sub> | β | P<sub>SGoF</sub> |
| SNPs not associated with AD individually |  |  |  |  |  |  |  |  |  |  |  |  |  |
| rs2018182 | rs12475670 | 0.048 | 1.00E+00 | 0.065 | 1.00E+00 | -0.179 | 1.31E-02 | 0.065 | 1.00E+00 | 0.048 | 1.00E+00 | -0.289 | 1.38E-02 |
| rs4143022 | rs35432737 | 0.020 | 1.00E+00 | -0.051 | 1.00E+00 | 0.248 | 2.02E-03 | -0.051 | 1.00E+00 | 0.020 | 1.00E+00 | 0.286 | 4.02E-02 |
| rs6431170 | rs55879780 | 0.142 | 2.84E-03 | 0.041 | 1.00E+00 | -0.014 | 1.00E+00 | 0.041 | 1.00E+00 | 0.142 | 2.84E-03 | -0.194 | 4.10E-02 |
| rs28387125 | rs62159566 | -0.125 | 4.86E-02 | -0.083 | 1.00E+00 | -0.004 | 1.00E+00 | -0.083 | 1.00E+00 | -0.125 | 4.87E-02 | 0.205 | 3.75E-02 |
| rs7598211 | rs4663046 | -0.081 | 1.37E-01 | -0.157 | 3.58E-03 | -0.043 | 1.00E+00 | -0.157 | 3.58E-03 | -0.081 | 1.37E-01 | 0.195 | 4.10E-02 |
| rs7562905 | rs55879780 | 0.115 | 5.54E-03 | 0.008 | 1.00E+00 | -0.102 | 1.00E+00 | 0.008 | 1.00E+00 | 0.115 | 5.54E-03 | -0.224 | 3.20E-02 |
| rs7599354 | rs7355740 | -0.256 | 2.58E-04 | 0.022 | 1.00E+00 | 0.055 | 1.00E+00 | 0.022 | 1.00E+00 | -0.256 | 2.58E-04 | 0.280 | 3.43E-02 |
| rs7599354 | rs13385611 | -0.147 | 5.29E-02 | 0.020 | 1.00E+00 | 0.157 | 6.38E-03 | 0.020 | 1.00E+00 | -0.147 | 5.34E-02 | 0.277 | 1.85E-02 |
| rs58478277 | rs10177866 | -0.035 | 1.00E+00 | -0.083 | 1.00E+00 | 0.139 | 8.07E-03 | -0.083 | 1.00E+00 | -0.035 | 1.00E+00 | 0.259 | 1.72E-02 |
| rs12478685 | rs72843819 | 0.176 | 2.06E-03 | 0.040 | 1.00E+00 | -0.115 | 1.00E+00 | 0.040 | 1.00E+00 | 0.176 | 2.06E-03 | -0.340 | 1.28E-02 |
| rs13016472 | rs7609186 | 0.171 | 2.30E-02 | 0.038 | 1.00E+00 | -0.111 | 1.00E+00 | 0.038 | 1.00E+00 | 0.171 | 2.30E-02 | -0.328 | 3.69E-02 |
| rs4663046 | rs4405698 | -0.111 | 5.57E-02 | -0.156 | 2.71E-02 | -0.051 | 1.00E+00 | -0.156 | 2.71E-02 | -0.111 | 5.61E-02 | 0.216 | 4.98E-02 |
| rs17014381 | rs7609186 | -0.178 | 5.10E-02 | 0.019 | 1.00E+00 | 0.192 | 6.10E-03 | 0.019 | 1.00E+00 | -0.178 | 5.10E-02 | 0.347 | 1.63E-02 |
| rs7597291 | rs7355740 | -0.149 | 2.50E-03 | -0.040 | 1.00E+00 | 0.129 | 1.00E+00 | -0.040 | 1.00E+00 | -0.149 | 2.50E-03 | 0.306 | 5.00E-02 |
| rs12619998 | rs7609186 | -0.459 | 2.71E-03 | -0.065 | 1.00E+00 | 0.042 | 1.00E+00 | -0.065 | 1.00E+00 | -0.459 | 2.71E-03 | 0.573 | 1.29E-02 |
| rs10167565 | rs7609186 | -0.368 | 1.69E-03 | -0.046 | 1.00E+00 | 0.116 | 1.00E+00 | -0.046 | 1.00E+00 | -0.368 | 1.69E-03 | 0.531 | 1.22E-02 |
| rs13419545 | rs12989081 | -0.156 | 1.73E-03 | -0.120 | 1.70E-02 | -0.081 | 1.00E+00 | -0.120 | 1.69E-02 | -0.156 | 1.73E-03 | 0.195 | 3.58E-02 |
| rs12466070 | rs61103548 | 0.053 | 1.00E+00 | 0.017 | 1.00E+00 | -0.209 | 8.98E-03 | 0.017 | 1.00E+00 | 0.053 | 1.00E+00 | -0.285 | 4.98E-02 |
| rs10928992 | rs2077309 | -0.086 | 1.00E+00 | -0.126 | 2.80E-02 | -0.015 | 1.00E+00 | -0.126 | 2.80E-02 | -0.086 | 1.00E+00 | 0.196 | 5.00E-02 |
| rs79801793 | rs6736826 | -0.118 | 6.99E-02 | -0.146 | 1.15E-02 | -0.008 | 1.00E+00 | -0.146 | 1.09E-02 | -0.118 | 7.00E-02 | 0.259 | 4.81E-02 |
| rs56279750 | rs12987692 | 0.056 | 1.00E+00 | 0.155 | 4.03E-04 | 0.025 | 1.00E+00 | 0.155 | 4.03E-04 | 0.056 | 1.00E+00 | -0.187 | 3.74E-02 |
| rs61146291 | rs72843819 | -0.057 | 1.00E+00 | -0.086 | 1.00E+00 | 0.172 | 9.81E-03 | -0.086 | 1.00E+00 | -0.057 | 1.00E+00 | 0.320 | 1.47E-02 |
| rs6710752 | rs7355740 | -0.209 | 2.32E-04 | -0.007 | 1.00E+00 | 0.091 | 1.00E+00 | -0.007 | 1.00E+00 | -0.209 | 2.32E-04 | 0.301 | 1.35E-02 |
| rs2276574 | rs7355740 | -0.218 | 1.22E-04 | 0.012 | 1.00E+00 | 0.130 | 1.00E+00 | 0.012 | 1.00E+00 | -0.218 | 1.22E-04 | 0.327 | 1.27E-02 |
| rs6759089 | rs7355740 | -0.309 | 2.17E-04 | -0.018 | 1.00E+00 | -0.015 | 1.00E+00 | -0.018 | 1.00E+00 | -0.309 | 2.17E-04 | 0.305 | 3.57E-02 |
| rs6759089 | rs13385611 | -0.185 | 1.08E-02 | -0.038 | 1.00E+00 | 0.104 | 1.00E+00 | -0.038 | 1.00E+00 | -0.185 | 1.08E-02 | 0.321 | 1.25E-02 |
| SNP(s) associated with AD individually |  |  |  |  |  |  |  |  |  |  |  |  |  |
| rs13398585* | rs11693422 | -0.041 | 1.00E+00 | -0.075 | 1.00E+00 | 0.086 | 1.00E+00 | -0.075 | 1.00E+00 | -0.041 | 1.00E+00 | 0.200 | 3.60E-02 |
| rs7609186 | rs6754293 | -0.092 | 7.19E-03 | 0.201 | 1.02E-02 | -0.216 | 4.05E-03 | 0.201 | 1.02E-02 | -0.092 | 7.19E-03 | -0.324 | 3.61E-02 |
| rs7609186 | rs58682665 | -0.147 | 1.72E-05 | 0.154 | 8.69E-02 | -0.318 | 1.67E-04 | 0.154 | 8.65E-02 | -0.147 | 1.72E-05 | -0.318 | 4.98E-02 |
| rs72843819 | rs6761391 | 0.150 | 1.14E-04 | 0.236 | 2.32E-03 | 0.120 | 1.00E+00 | 0.236 | 2.32E-03 | 0.150 | 1.14E-04 | -0.274 | 4.83E-02 |
| rs934827 | rs6761391 | 0.194 | 4.30E-05 | 0.135 | 7.95E-02 | 0.107 | 6.58E-02 | 0.135 | 7.94E-02 | 0.194 | 4.30E-05 | -0.223 | 3.20E-02 |
| rs11693422 | rs7584040 | 0.150 | 7.38E-04 | 0.174 | 6.95E-05 | 0.114 | 1.25E-02 | 0.174 | 6.95E-05 | 0.150 | 7.38E-04 | -0.211 | 1.37E-02 |
| rs7575209 | rs2077309 | -0.036 | 1.00E+00 | -0.247 | 6.68E-06 | -0.059 | 1.00E+00 | -0.247 | 6.68E-06 | -0.036 | 1.00E+00 | 0.223 | 1.29E-02 |
| rs58682665 | rs2253005 | -0.108 | 1.56E-02 | -0.282 | 3.37E-08 | -0.203 | 7.47E-06 | -0.282 | 3.37E-08 | -0.108 | 1.56E-02 | 0.186 | 4.00E-02 |

Notations: AD = Alzheimer's Disease; SNP = single-nucleotide polymorphism; Int = interaction term; CompG = compound genotype; mM (or Mm) = heterozygous CompG comprised of one or two minor alleles of SNP1 (or SNP2) and two major alleles of the other SNP; mm = homozygous CompG composed of one or two minor alleles of each SNP1 and SNP2; $\beta$ = CompG effect in the CompG models or SNP1, SNP2, and interaction effects in the interaction models; P<sub>SGoF</sub> = SGoF-adjusted P-value; SGoF = Sequential Goodness-of-Fit. \*The rs13398585-rs11693422 pair had significant mm-mM difference with P<sub>SGoF</sub><2.04E-02, although its Mm, mM, and mm effects were not significant.

**Table 4.** Ten AD-associated SNP pairs mapped to the *MSA4A6* gene locus with significant CompG effects and SNP-by-SNP interactions.

| SNP Pair |  | Compound Genotype Model |  |  |  |  |  | Interaction Model |  |  |  |  |  |
| --- | --- | --- | --- | --- | --- | --- | --- | --- | --- | --- | --- | --- | --- |
| SNP1 | SNP2 | Mm |  | mM |  | mm |  | SNP1 |  | SNP2 |  | Int |  |
| | | $\beta$ | P<sub>SGoF</sub> | $\beta$ | P<sub>SGoF</sub> | $\beta$ | P<sub>SGoF</sub> | $\beta$ | P<sub>SGoF</sub> | $\beta$ | P<sub>SGoF</sub> | $\beta$ | P<sub>SGoF</sub> |
| SNPs not associated with AD individually |  |  |  |  |  |  |  |  |  |  |  |  |  |
| rs570322 | rs11230137 | 0.109 | 5.76E-03 | 0.112 | 9.10E-02 | 0.003 | 1.00E+00 | 0.112 | 8.91E-02 | 0.109 | 5.79E-03 | -0.219 | 2.06E-02 |
| rs56162497 | rs117265279 | -0.057 | 1.00E+00 | -0.079 | 1.00E+00 | 0.281 | 5.07E-03 | -0.079 | 1.00E+00 | -0.057 | 1.00E+00 | 0.419 | 1.99E-02 |
| rs141802649 | rs10897068 | -0.031 | 1.00E+00 | -0.061 | 1.00E+00 | 0.148 | 2.95E-02 | -0.061 | 1.00E+00 | -0.031 | 1.00E+00 | 0.238 | 4.31E-02 |
| rs1286386 | rs4939392 | 0.160 | 4.24E-02 | 0.196 | 9.77E-04 | 0.145 | 1.09E-02 | 0.196 | 9.77E-04 | 0.160 | 4.26E-02 | -0.211 | 2.03E-02 |
| rs1286383 | rs11230137 | 0.211 | 4.36E-03 | 0.114 | 1.01E-02 | 0.105 | 1.36E-02 | 0.114 | 1.01E-02 | 0.211 | 4.36E-03 | -0.222 | 2.04E-02 |
| rs1286383 | rs117265279 | -0.244 | 2.53E-03 | 0.006 | 1.00E+00 | 0.114 | 5.73E-02 | 0.006 | 1.00E+00 | -0.244 | 2.53E-03 | 0.356 | 1.96E-02 |
| rs11230137 | rs117265279 | -0.087 | 1.00E+00 | 0.020 | 1.00E+00 | 0.159 | 1.30E-02 | 0.020 | 1.00E+00 | -0.087 | 1.00E+00 | 0.228 | 3.63E-02 |
| rs2583477 | rs117265279 | -0.234 | 1.21E-02 | 0.037 | 1.00E+00 | 0.115 | 4.72E-02 | 0.037 | 1.00E+00 | -0.234 | 1.23E-02 | 0.316 | 2.03E-02 |
| rs1786140 | rs2000636 | -0.149 | 4.26E-02 | -0.036 | 1.00E+00 | 0.010 | 1.00E+00 | -0.036 | 1.00E+00 | -0.149 | 4.35E-02 | 0.195 | 2.19E-02 |
| SNP(s) associated with AD individually |  |  |  |  |  |  |  |  |  |  |  |  |  |
| rs17154321 | rs1286170 | -0.221 | 2.08E-05 | -0.087 | 1.00E+00 | -0.130 | 5.27E-03 | -0.087 | 1.00E+00 | -0.221 | 2.08E-05 | 0.178 | 2.27E-02 |

Notations: AD = Alzheimer's Disease; SNP = single-nucleotide polymorphism; Int = interaction term; CompG = compound genotype; mM (or Mm) = heterozygous CompG comprised of one or two minor alleles of SNP1 (or SNP2) and two major alleles of the other SNP; mm = homozygous CompG composed of one or two minor alleles of each SNP1 and SNP2; $\beta$ = CompG effect in the CompG models or SNP1, SNP2, and interaction effects in the interaction models; P<sub>SGoF</sub> = SGoF-adjusted P-value; SGoF = Sequential Goodness-of-Fit.

### 3.1 *BIN1* gene results

Our single SNP meta-analyses showed that 18 of 139 SNPs of interest in the *BIN1* locus were associated with AD at P_SGoF_<0.05 (Table 2), with minor alleles of 12 of them having positive effects and those of the other six having negative effects on the AD risk. The most significant AD-associated *BIN1* SNP was rs4663105 with β=0.239, P=1.51E-13, and P_SGoF_ =8.25E-04. For these 18 SNPs, the pairwise LD magnitudes measured by |r| were between 0.001 and 0.826 in the AD-affected group and between 0.002 and 0.819 in the AD-unaffected group. These 18 SNPs were mostly in low LD with each other. Less than one-third of the 153 pairs constructed from them had |r|≥0.316 (i.e., r^2^≥0.1) and only six of these pairs had |r|≥0.7 in each group (Table S5).

Among 9591 *BIN1* SNP pairs in our CompG meta-analyses, 589 pairs showed significant CompG effects at P_SGoF_<0.05 (Table S3 and Figure 1), whereas neither of the SNPs in these pairs were individually associated with AD in single SNP models. These included 213, 97, and 242 pairs with significant Mm, mM, and mm effects, respectively, and 37 pairs with more than one significant CompG effects.

**Figure 1.**
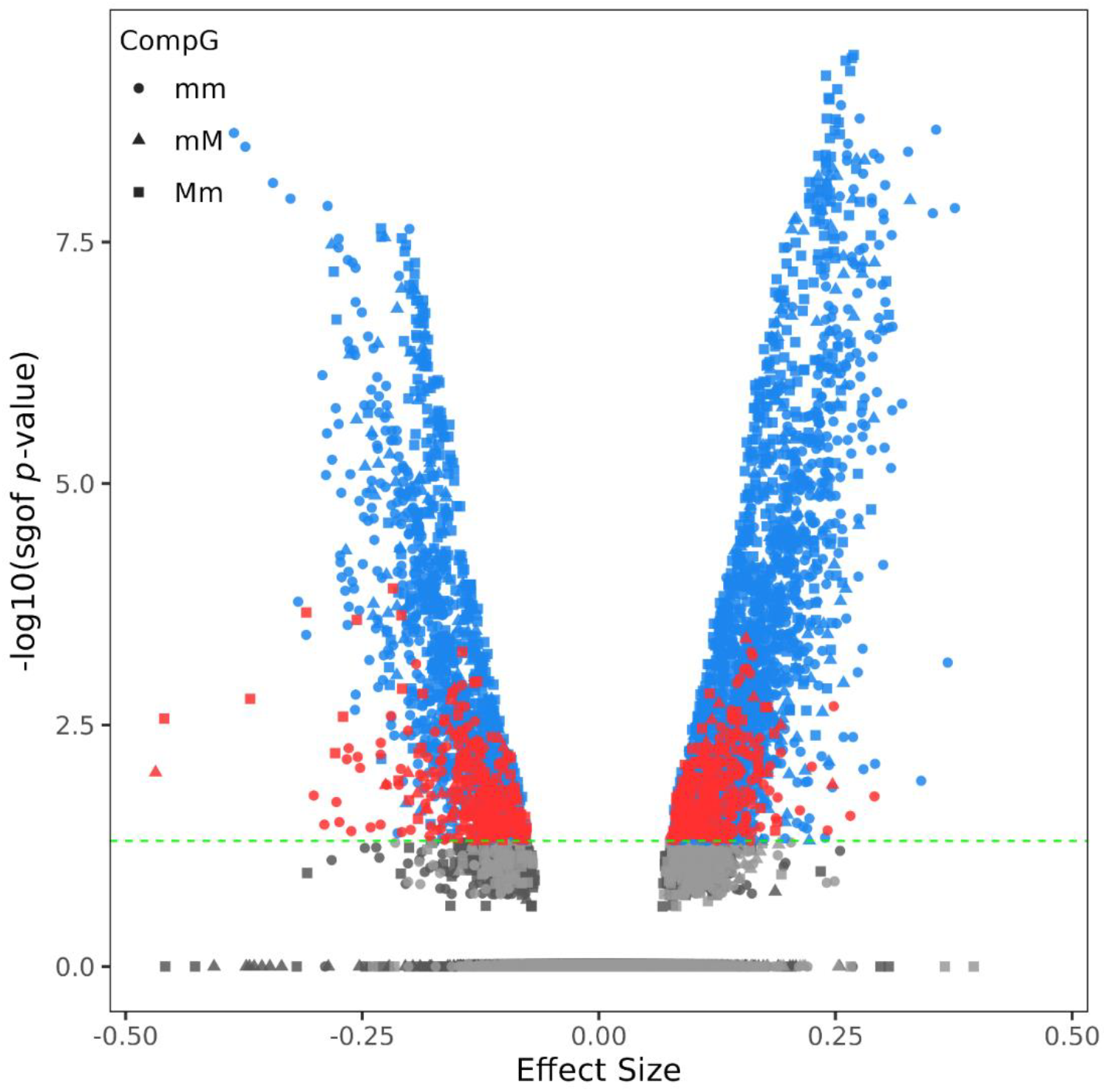
Volcano plot for CompG analyses of the 9591 SNP pairs mapped to the *BIN1* gene locus on chromosome 2q14.3. The x-axis displays CompG regression beta coefficients, and the y-axis shows the respective minus-logarithm-base-10 of SGOF-adjusted P-values. The dashed line indicates the significance threshold [i.e., −log_10_(P_SGoF_=0.05)=1.3]. The red dots above the cutoff line indicate CompG effects whose comprising SNPs were not associated with Alzheimer’s disease (AD) individually. The blue dots above the cutoff line denote significant CompG effects, in which one or both comprising SNPs were associated with AD individually. The light-gray or dark-gray dots below the cutoff line show non-significant CompG effects, in which none or at least one of the comprising SNPs were associated with AD individually.

Additionally, 38 SNP pairs, whose comprising SNPs as well as their Mm, mM, and mm effects were not associated with AD, had significant mm-mM (12 pairs) and/or Mm-mM (27 pairs) differences in the chi-square test (Table S3 and Figure 1). For six of the 27 SNP pairs with significant Mm-mM differences (i.e., rs9646710-rs35322174, rs35369712-rs72843819, rs74598592-rs6708915, rs56279750-rs61103548, rs544587-rs61103548, and rs4663096-rs113528216), the P-values for Mm-mM chi-square differences (1.69E-02, 1.18E-02, 1.57E-02, 1.17E-02, 1.21E-02, and 1.74E-02, respectively) were smaller than those obtained from chi-square test for the differences in the effects of their comprising SNPs in the single SNP models (3.14E-02, 1.44E-02, 4.47E-02, 4.31E-02, 4.86E-02, and 4.94E-02, respectively). This may indicate a partial interaction between the SNPs in each pair, as the minor allele effect of one SNP became more significant in the absence of the minor allele of the other SNP.

Of note, 26 of these 627 (i.e., 589+38) SNP pairs had significant SNPxSNP interaction terms (P_SGoF_<0.05) in the traditional interaction models (Tables 3 and S3). The LD magnitudes for these 26 SNP pairs were relatively small, with |r| coefficients ranging from 0.002 to 0.435 in the cases and from 0.001 to 0.411 in the controls (Tables S5 and S6).

The LD magnitudes (i.e., |r|) for these 627 SNP pairs were between 1.80E-05 and 0.835 in the cases and between 1.86E-04 and 0.830 in the controls. SNPs in most pairs were in low LD, with only 23 and 22 pairs having |r|≥0.316 (i.e., r^2^≥0.1) in the two groups, respectively. Only four pairs in each group had |r|≥0.7. The chi-square test comparing LD r coefficients of these 627 SNP pairs between the AD-affected and AD-unaffected groups identified four pairs with significantly different LD in the two groups at Bonferroni-adjusted significance level P<7.97E-05 (=0.05/627). All four pairs had a larger |r| in the AD-affected group (Tables S5 and S6).

Our CompG meta-analyses also revealed 1967 additional AD-associated CompGs at P_SGoF_<0.05, in which one or both comprising SNPs were individually associated with AD as well (Table S4 and Figure 1). These included 419, 215, and 292 pairs with significant Mm, mM, and mm effects, respectively, as well as 1041 pairs with more than one significant CompG effects. When the chi-square test was used to compare significant effects from CompG models with those from the single SNP models, no significant P-values were obtained at Bonferroni-adjusted significance level P<2.54E-05 (=0.05/1967), indicating that the significant CompG effects could mainly be attributed to the corresponding significant SNP main effects.

Additionally, 76 SNP pairs with at least one SNP in a pair individually associated with AD but showing no significant effects for Mm, mM, and mm, had significant mm-mM (14 pairs) and/or Mm-mM (72 pairs) differences in the chi-square test (Table S4 and Figure 1). For one of the 72 SNP pairs with significant Mm-mM differences (rs745717-rs113528216), the Mm-mM P-value (P=4.79E-03) was smaller than the chi-square test P-value (P=6.94E-03) comparing the main effects of the two SNPs in single SNP models. This could imply the partial interaction of the two SNPs, in which the effect of minor allele of rs113528216 became more prominent in the absence of the minor allele of rs745717 (i.e., Mm genotype carriers).

Among these 2043 (i.e., 1967+76) SNP pairs, eight pairs had significant SNPxSNP interaction terms (P_SGoF_<0.05) in the traditional interaction models (Tables 3 and S4). The |r| coefficients for these eight SNP pairs were between 0.003 and 0.114 in the AD-affected group and between 0.019 and 0.095 in the AD-unaffected group, highlighting their low LD (Tables S5 and S6).

The LD |r| values for these 2043 SNP pairs ranged from 5.03E-06 to 0.826 in the case group and from 2.44E-05 to 0.819 in the control group, with 85 and 80 pairs having |r|≥0.316 (i.e., r^2^≥0.1) in the two groups, respectively. Only seven and eight pairs had |r|≥0.7 in the two groups, respectively. When the LD r coefficients for each of these 2043 SNP pairs were compared between the two groups through chi-square test, 11 pairs had significantly different LD in the AD-affected and AD-unaffected groups at Bonferroni-adjusted significance level P<2.44E-05 (=0.05/2043). Six of them had a larger |r| while the other five pairs had a smaller |r| in the AD-affected group (Tables S5 and S6).

#### 3.2 *MS4A6A* gene results

Four of 66 *MS4A6A* SNPs of interest were associated with AD at P_SGoF_<0.05 in the single SNP meta-analyses (Table 2). The minor alleles of all SNPs had negative effects on the AD risk, with rs1286289 having the most significant signal (β=-0.151, P=4.21E-07, and P_SGoF_ =8.75E-03). The LD magnitudes measured by |r| for the six SNP pairs constructed from these four SNPs were relatively modest and ranged from 0.358 to 0.816 in the case group and from 0.364 to 0.829 in the control group. Only one pair had |r|≥0.316 (i.e., r^2^≥0.1) in each group (Table S9).

Among 2145 *MS4A6A* SNP pairs of interest, 217 pairs had significant CompG effects at P_SGoF_<0.05 in our AD-CompG meta-analyses, while none of their comprising SNPs were individually associated with AD in the single SNP analyses. These included 40, 48, and 87 pairs with significant Mm, mM, and mm effects, respectively, along with 42 pairs, which had more than one significant CompG effects (Table S7 and Figure 2).

**Figure 2.**
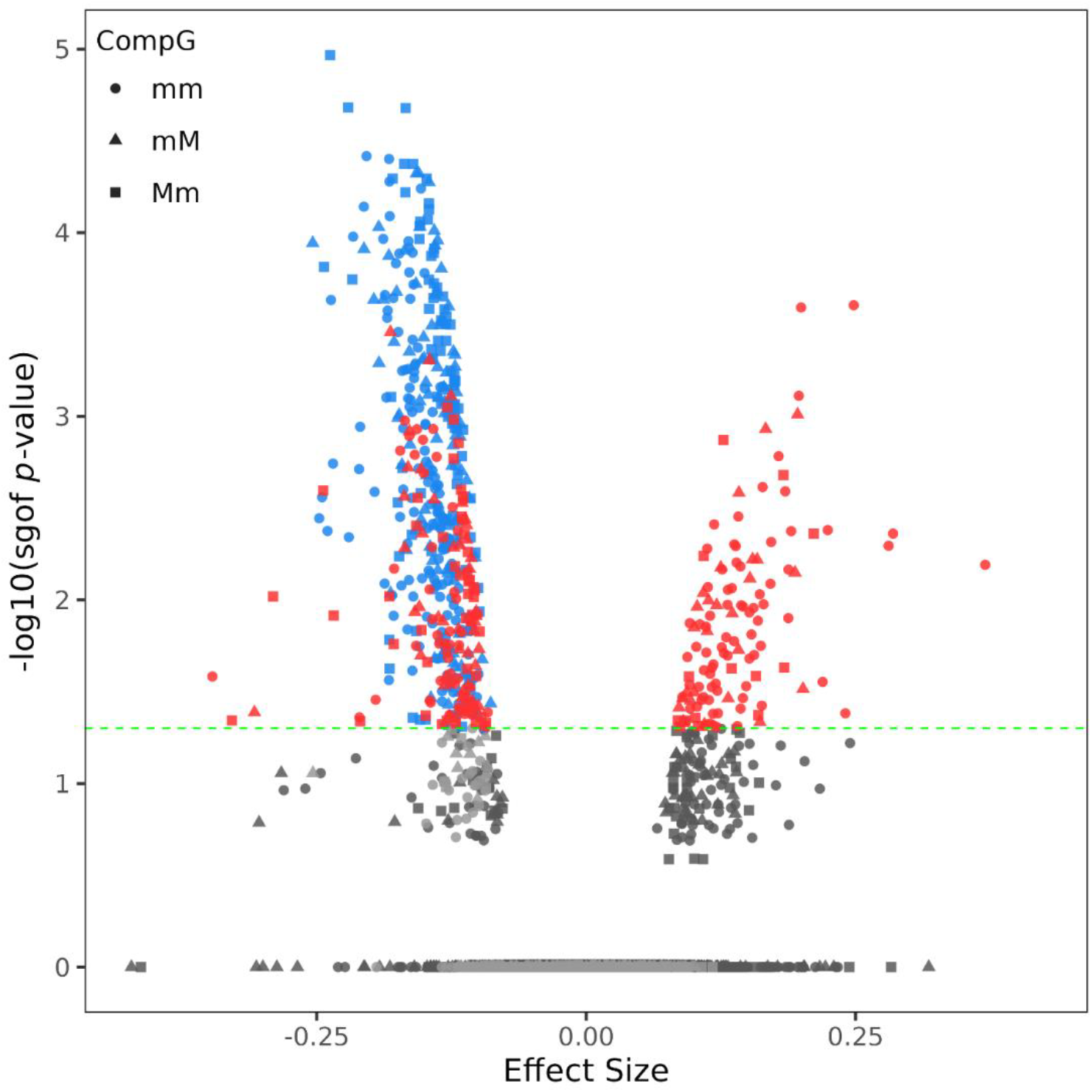
Volcano plot for CompG analyses of the 2145 SNP pairs mapped to the *MS4A6A* gene locus on chromosome 11q12.2. The x-axis displays CompG regression beta coefficients, and the y-axis shows the respective minus-logarithm-base-10 of SGOF-adjusted P-values. The dashed line indicates the significance threshold [i.e., −log_10_(P_SGoF_=0.05)=1.3]. The red dots above the cutoff line indicate CompG effects whose comprising SNPs were not associated with Alzheimer’s disease (AD) individually. The blue dots above the cutoff line denote significant CompG effects, in which one or both comprising SNPs were associated with AD individually. The light-gray or dark-gray dots below the cutoff line show non-significant CompG effects, in which none or at least one of the comprising SNPs were associated with AD individually.

There were four additional SNP pairs, whose comprising SNPs and Mm, mM, and mm effects were not associated with AD, although their mm-Mm (one pair) or Mm-mM (three pairs) differences were significant in the chi-square test (Table S7 and Figure 2).

Only nine of these 221 (i.e., 217+4) SNP pairs had significant (P_SGoF_<0.05) SNPxSNP interaction terms in traditional interaction models (Tables 4 and S7). The LD magnitudes for these nine SNP pairs were small (i.e., |r| coefficients between 0.005 and 0.322 in the AD affected group and between 0.015 and 0.353 in the AD unaffected group) (Tables S9 and S10).

These 221 SNP pairs had LD magnitudes (i.e., |r| values) of 5.30E-04 to 0.784 in the AD-affected group and 5.67E-05 to 0.809 in the AD-unaffected group. Once again, SNPs in most pairs were in low LD as only 16 and two pairs had |r|≥0.316 (i.e., r^2^≥0.1) and |r|≥0.7, respectively, in each group. When the LD r coefficients of each of these 221 SNP pairs were compared between the case and control groups by a chi-square test, 18 pairs were found to have significantly different LD in the two groups at Bonferroni-adjusted significance level P<2.26E-04 (=0.05/221). Of these, 10 pairs had a larger |r| and eight pairs had a smaller |r| in the AD-affected group (Tables S9 and S10).

Additionally, our AD-CompG meta-analyses identified 237 SNP pairs, which had significant CompG effects (P_SGoF_<0.05), while at least one SNP in each pair was also associated with AD individually in the single SNP models (Table S8 and Figure 2). These included 43 pairs with significant Mm effects, 37 pairs with significant mM effects, and 51 pairs with significant mm effects, and 106 pairs with more than one significant CompG effects. As expected, the significant CompG effects were primarily due to their counterpart significant SNP main effects in the single SNP models as the CompG effects were not significantly different from SNP main effects at Bonferroni-adjusted significance level P<2.11E-04 (=0.05/237) in the chi-square test.

There were five additional SNP pairs, which had non-significant Mm, mM, and mm effects, but significant Mm-mM differences in the chi-square test (Table S8 and Figure 2).

Only one of these 242 (i.e., 237+5) SNP pairs had significant SNPxSNP interaction term (P_SGoF_<0.05) in the traditional interaction models (Tables 4 and S8). The LD |r| coefficients for this pair were 0.114 and 0.126 in the AD-affected and AD-unaffected groups, respectively (Tables S9 and S10).

For these 242 SNP pairs, the LD magnitudes measured by |r| were between 6.96E-04 and 0.816 in cases and between 0.001 and 0.829 in controls, with 25 and three pairs having |r|≥0.316 (i.e., r^2^≥0.1) and |r|≥0.7, respectively, in each group. The chi-square test comparing LD r coefficients between these two groups showed that six of these 242 pairs had significantly different LD in the AD-affected and AD-unaffected groups at Bonferroni-adjusted significance level P<2.07E-04 (=0.05/242) [two pairs with larger and four pairs with smaller LD magnitudes in the AD-affected group] (Tables S9 and S10).

## 4. Discussion

We investigated the associations of the AD risk with two sets of 139 and 66 SNPs (and their respective 9591 and 2145 SNP pairs) mapped to the *BIN1* and *MS4A6A* loci on chromosomes 2q14.3 and 11q12.2, respectively, leveraging three dominant allelic-effect models including single SNP, compound genotypes, and traditional SNPxSNP interaction models. These analyses identified novel AD associations at both individual SNP and SNP-pair levels.

We found that 18 *BIN1* SNPs and 4 *MS4A6A* SNPs were individually associated with AD with relatively small effect sizes. While the minor alleles of all four *MS4A6A* SNPs showed protective associations with AD, the minor alleles of most of the identified *BIN1* SNPs (12 of 18) demonstrated adverse effects. The pairwise LD magnitudes in each of these two SNP sets were mostly small (r^2^<0.1). Previous GWAS have reported the AD associations of seven of these *BIN1* SNPs [10], including rs11680911 [58], rs4663105 [14,21,27,59–62], rs35103166, rs58682665, rs730482, rs745717, and rs7575209 [20]. The minor alleles of all seven but two SNPs (i.e., rs58682665 and rs7575209) were adversely (i.e., positive betas) associated with AD in our single SNP analyses.

Our analyses of associations of the 9591 and 2145 SNP pairs mapped to the *BIN1* and *MS4A6A* loci with AD provided the following main insights. First: We found that there were 1967 *BIN1* and 237 *MS4A6A* SNP pairs whose CompG (i.e., combinations of genotypes) effects as well as one or both of their comprising SNPs were associated with AD. The significant CompG effects in such SNP pairs are likely driven by their significant individual SNP main effects.

Second: There were two smaller sets of 589 and 217 AD-associated SNP pairs mapped to the *BIN1* and *MS4A6A* loci, respectively, in which none of the SNPs were associated with AD in the single SNP models. The SNPs in most of these pairs were in low LD, with only ∼3.5% and ∼7% of these SNP pairs having r^2^≥0.1, respectively.

Among the 589 *BIN1* SNP pairs, significant effects were observed for heterozygous CompG (i.e., Mm and/or mM) from 316 pairs, homozygous CompG (i.e., mm) from 242 pairs, and both heterozygous and homozygous CompGs from 31 pairs. The significant CompGs had relatively small effect sizes, ranging from -0.468 to 0.291, and most of them (55%) were adversely associated with AD. Also, 401 of these 589 SNP pairs, including 95.6% and 49% of the pairs with significant effects for homozygous and heterozygous CompG, respectively, were comprised of SNPs with the same directions of effects in the single SNP models.

Among the 217 *MS4A6A* SNP pairs, significant effects were identified for heterozygous CompG from 89 pairs and homozygous CompG from 87 pairs. The other 41 pairs had significant effects for both heterozygous and homozygous CompGs. Again, most of them (56%) were associated with increased AD risk and generally had small effect sizes, ranging from -0.347 to 0.370 (except for one pair, i.e., rs2194961-rs7117320, with Mm effect of 2.09, which might be due to the small sample size of Mm genotype for this pair). The SNPs comprising 145 of these 217 SNP pairs had the same directions of effects in the single SNP models. These included 90.6% and 48.5% of SNP pairs with significant effects for homozygous and heterozygous CompGs, respectively.

Third: There were seven additional *BIN1* SNP pairs whose CompG effects were not associated with AD directly, however, the differences for their heterozygous CompGs (i.e., Mm-mM) were more prominent than the differences of their individual SNP main effects. These can be due to partial interaction between the SNPs in each pair, where the effect of the minor allele of one SNP became more prominent when the minor allele of the other SNP is not present.

Fourth: Notably, 34 and 10 SNP pairs mapped to the *BIN1* and *MS4A6A* genes, respectively, had significant interaction terms in the traditional SNPxSNP interaction models, in addition to the significant CompG effects. These SNP pairs were mostly independent due to their weak pairwise LD. Only three *BIN1* SNP pairs and one *MS4A6A* SNP pair had |r|≥0.316 (i.e., r^2^≥0.1) and none of them had |r|≥0.7. For 31 of 34 *BIN1* and 9 of 10 *MS4A6A* SNP pairs, the effect sizes of the interaction terms were larger than those of the main effects of the comprising SNPs. Also, for 22 and 7 of these SNP pairs, respectively, the interaction terms were positively associated with AD. The findings from our CompG analyses and interaction models were not fully overlapping, i.e., the vast majority of the SNP pairs with significant CompG effects did not have significant SNPxSNP interaction terms, and, in addition, only 11 *BIN1* and 7 *MS4A6A* SNP pairs had both significant interaction terms and compound homozygous CompG (i.e., mm). Therefore, implementing both CompG and traditional interaction analyses is helpful for comprehensively exploring the genetic architecture of AD and other complex traits [43].

The identified significant CompG and interaction effects support previous findings on potential roles of *BIN1* and *MS4A6A* loci in AD development [11–21,40,59,60,62–64], and illustrate the complex genetic landscape of AD within these loci, which contributes to the genetic heterogeneity of AD. Prior studies provided some insights into complex AD-related associations within these two loci [18,40,59,63,64]. For instance, the *APOE* ε4 allele was found to interact with *BIN1* rs744373 and *MS4A6A* rs610932 SNPs modifying their effects on the short-term memory scores [64]. Also, interactions between *MS4A4E/6A* rs670139 SNP and SNPs in the other AD-risk genes like *CLU* rs11136000 SNP and *CD33* rs3865444 SNP have been reported at the nominal significance level with P<0.003 and P<0.016, respectively [63]. Of these, the *APOE* ε4 allele could modify the rs670139-by-rs11136000 epistatic effect on the AD risk [40]. Previous reports have suggested that the association signals in the *MS4A6A* locus with AD were stronger in the *APOE* ε4 negative sample [18,40,59]. The significant findings from our CompG and interaction analyses expand the current knowledge regarding complex genetic associations in these two loci. Similarly, such analyses have resulted in significant findings in the other AD-associated loci. For instance, CompG analyses have identified ε2-independent association of rs2884183 (*DDX10* variant) and ε4-independent association of rs483082 (*APOC1* variant) with AD [39]. Also, several SNPs not individually associated with AD were found to form SNP pairs with AD-associated CompG effects in the *CLU* and *ABCA7* loci [43].

Among SNP pairs with significant findings in our current CompG analyses, 15 *BIN1* and 24 *MS4A6A* SNP pairs had significantly different LD r coefficients in the AD-affected and AD-unaffected groups, with LD magnitudes (i.e., |r|) for 10 and 12 of such SNP pairs, respectively, being larger in the AD-affected group. Such LD differences have been previously examined in other AD-risk genes. In particular, larger and smaller magnitudes of LD of SNPs in the *APOE* 19q13.3 locus with rs429358 (*APOE* ε4-encoding SNP) and rs7412 (*APOE* ε2-encoding SNP), respectively, were identified as important molecular characteristics of AD [37,65].

## 5. Conclusions

The results presented here provide novel insights into the contributions of the *BIN1* and *MS4A6A* loci to the heterogenous genetic architecture of AD. These included individual SNP and complex nonadditive (in the forms of CompGs, SNPxSNP interactions, and partial SNP interactions in compound heterozygotes) associations with AD. In particular, 589 *BIN1* and 217 *MS4A6A* SNP pairs with AD-associated CompGs whose comprising SNPs were not associated with AD individually could indicate important sources of genetic heterogeneity of AD in these loci. Notably, two sets of 34 and 10 SNP pairs in these loci, respectively, had both significant SNPxSNP interaction terms and CompG effects. Also, seven other *BIN1* SNP pairs showed evidence of partial interactions, wherein differences in their heterozygous CompGs could not be fully explained by the differences of their individual SNP main effects. Taken together, these findings emphasize the importance of exploring higher-level genetic associations beyond the individual SNP level when investigating the genetic architecture of AD.

## Data Availability

The four analyzed datasets are available to the qualified researchers by the dbGaP (ADGC, ADSP, and LOAD-FBS datasets), NIAGADS (ADSP dataset), and the UK Biobank (UKB dataset).

https://www.ncbi.nlm.nih.gov/gap

https://www.niagads.org/adsp/content/home

https://www.ukbiobank.ac.uk

## Statements and Declarations

## Acknowledgments

This study used limited access data from dbGaP [accession numbers: phs000372.v1.p1 (ADGC), phs000572.v8.p4 (ADSP), and phs000168.v2.p2 (LOAD-FBS)], NIAGADS [accession number: NG00067 (ADSP-WGS)], and the UK Biobank [applications numbers: 60447 and 62778 (UKB)]. Please see detailed *Supporting Acknowledgment* in the Supplementary Information File regarding the four analyzed datasets.

## Ethics Approval and Consent to Participate

This study does not involve gathering data from human subjects directly. Instead, it focuses on secondary analysis of data obtained from dbGaP, NIAGADS, and the UK Biobank. The data were accessed with the approval of the Duke Health Institutional Review Board (IRB) [protocols: Pro00105245-INIT-1.0 (06/26/2020), Pro00105247-INIT-1.0 (06/26/2020), and Pro00105346-INIT-1.0 (04/15/2020)], and all analyses were performed under IRB guidelines.

## Availability of Data and Materials

The four analyzed datasets are available to the qualified researchers by the dbGaP (https://www.ncbi.nlm.nih.gov/gap), NIAGADS (https://www.niagads.org/adsp/content/home), and the UK Biobank (https://www.ukbiobank.ac.uk).

## Supplementary Materials

1) Supplementary Information File containing *Supporting Acknowledgment*, and 2) Tables S1-S10 in Excel format.

